# Sudden cardiac death in the young: A qualitative study of experiences of family members with cardiogenetic evaluation

**DOI:** 10.1101/2022.09.28.22280480

**Authors:** Lieke van den Heuvel, Judy Do, Laura Yeates, Charlotte Burns, Chris Semsarian, Jodie Ingles

**Affiliations:** Cardio Genomics Program at Centenary Institute, The University of Sydney, Sydney, Australia; Department of Clinical Genetics, Amsterdam UMC (location AMC), Amsterdam, the Netherlands; Department of Genetics, University Medical Centre Utrecht, Utrecht, the Netherlands; Netherlands Heart Institute, Utrecht, the Netherlands; Centre for Population Genomics, Garvan Institute of Medical Research, and UNSW Sydney, Sydney, Australia; Centre for Population Genomics, Murdoch Children’s Research Institute, Melbourne, Australia; Agnes Ginges Centre for Molecular Cardiology at Centenary Institute, The University of Sydney, Sydney, Australia; Faculty of Medicine and Health, The University of Sydney, Sydney, Australia; Department of Cardiology, Royal Prince Alfred Hospital, Sydney, Australia

**Keywords:** Sudden cardiac death, cardiogenetic evaluation, postmortem genetic testing, experiences, perceptions, family members

## Abstract

**Introduction:** Sudden cardiac death (SCD) is a devastating event for the family and the community, especially when it occurs in a young person (<45 years). Genetic heart diseases, including cardiomyopathies and primary arrhythmia syndromes, are an important cause of SCD in the young. Although cardiogenetic evaluation, i.e., clinical evaluation, genetic testing and psychological support, is increasingly performed after SCD, it is unknown how suddenly bereaved family members experience the process. We aimed to explore the experiences of family members with cardiogenetic evaluation after SCD, and their perception of the process and care received.

**Methods:** In-depth interviews were conducted with eighteen family members of young people (<45 years old) who died suddenly, including parents, siblings and partners. The interviews were thematically analysed by two researchers independently.

**Results:** In total, 18 interviews were conducted from 17 families. The following themes were identified: (1) Experiences with postmortem genetic testing including managing expectations and psychological impact, (2) appreciation of care such as access to genetic counselling and relief following cardiac evaluation of relatives, and (3) need for support including unmet psychological support needs and better coordination of care immediately after the death.

**Conclusion:** Although participants appreciated the opportunity for cardiogenetic evaluation, they also experienced a lack of coordination of cardiogenetic and psychological care. Our findings stress the importance of access to expert multidisciplinary teams, including psychological care, to adequately support these families after a SCD in a young family member.

## INTRODUCTION

Sudden cardiac death (SCD) is a tragic event that is devastating for the family as well as the community, especially when it occurs in a young person (< 45 years). Genetic heart diseases are an important cause of death in such cases and can include inherited cardiomyopathies and primary arrhythmia syndromes (1). Most are inherited in an autosomal dominant pattern, meaning that families dealing with the profound grief of the death must also confront the heritable risk of disease for themselves and close family members.

Where a SCD is due to a primary arrhythmia syndrome, such as long QT syndrome or Brugada syndrome, no structural abnormalities will be identified at postmortem investigation. Postmortem genetic testing may identify an underlying genetic cause in approximately 10-20% (1-3). If a causative variant can be identified, then cascade genetic testing can be offered to at-risk family members, clarifying risk for relatives and guiding clinical evaluation. Importantly, for many families, identifying an underlying genetic cause can also provide an answer for the death, which may also help the grieving process. First-degree family members are at increased risk of complicated grief and posttraumatic stress symptoms, especially mothers of the deceased (4,5). Literature describing needs of these families indicate a desire for reconstruction of the event, a need for sensitivity from healthcare professionals involved in their care, and a desire for appropriate psychosocial care (6,7). Cardiogenetic evaluation incorporates the clinical, genetic and psychological investigations that should be offered to families after a young SCD (8).

Although postmortem genetic testing is increasingly performed after SCD, the experiences and perceptions of family members regarding cardiogenetic evaluation, including postmortem genetic testing, is largely unexplored. We have previously shown greater psychological distress have poor adaptation to their genetic results among a small group of relatives who had experienced a young SCD (9). Here we explore the experiences of family members with cardiogenetic evaluation after SCD, and their perception of the process and care received including the genetic test result. Insights gained could be used to improve the care for these families.

## METHODS

### Design

A qualitative interview study design was used to explore the experiences of family members of young people who died suddenly, with in-depth focus on cardiogenetic evaluation and other care they received following the SCD.

### Participants

Adult family members of young people who died suddenly (<45 years old), including parents, siblings and partners, attending a multidisciplinary specialised cardiogenetic clinic in Sydney, Australia, from 2003 to 2020 were eligible for inclusion in this study. Family members of a young person who died very recently (i.e., < 6 months) were not approached. A purposive sampling strategy was used, whereby we aimed to include families with a range of clinical and demographic characteristics.

### Procedures

Family members of SCD cases were approached by phone by a genetic counsellor involved in the care for these families (LY). If interested, an information sheet was provided via email. Written informed consent was requested. Semi-structured interviews were conducted by LvdH, a researcher with a background in psychology and not involved in the care of these families. Interviews were conducted over the phone or face-to-face, depending on the preferences of participants. All interviews were audio-recorded with consent of the participants. A semi-structured interview scheme was developed based on existing literature and clinical experience, and was reviewed by the multidisciplinary team. Topics covered were: (1) experiences with being informed about the possibility of postmortem genetic testing, (2) their appreciation of the care they received, (3) experiences with the process of postmortem genetic testing, (4) and perceptions of the result of postmortem genetic testing (see **Supplementary Material S1**). Recruitment of participants for the interviews was continued until data saturation was reached. This study was approved by the Sydney Local Health District (Royal Prince Alfred Hospital Zone) Ethics Committee (X19-0019 & 2019/ETH00094) and participants provided written consent.

### Data analysis

All audio-recordings were transcribed verbatim by an external transcriber. For one of the interviews, the quality of the recording was insufficient and was therefore not transcribed nor analysed. Thematic data analysis was conducted by two researchers (LvdH and JD) independently, based on the principles of Braun and Clarke (10). After initial coding of the interviews, a codebook was developed. Any disagreements between the two researchers were discussed until agreement was met. A third researcher (JI) was available for consultation in case the two researchers did not reach agreement. Based on the codebook, themes and subthemes were created and discussed amongst the author group. All transcripts were re-read by LvdH to ensure no interesting findings were missed. Quotes of interviews were collected to illustrate the results.

## RESULTS

### Participants

**Table 1** shows the sociodemographic and clinical characteristics of participants. In total, 18 family members from 17 families completed an interview. Interviews were conducted with 10 parents, 5 siblings and 3 partners of young people who suddenly died. In the family of 9 participants, a potential genetic disease was identified at postmortem examination, including inherited cardiomyopathies and arrhythmia syndromes. A disease-causing genetic variant was identified in 7 (41%) families. In the other families, results of postmortem genetic testing were considered uninformative, including variants of uncertain significance (VUS) (n=6) and where no variant was identified (n=5). Three families had their genetic variant reclassified, 2 downgraded and 1 upgraded.

**TABLE 1.** Sociodemographic and clinical characteristics of participants

|  | **N (%) or N ± SD** |
| --- | --- |
| *Sex* |  |
| Female | 11 (61.1) |
| Male | 7 (38.9) |
| Mean age (SD) | 49.3 ± 10.2 |
| Time since SCD in years | 8.1 ± 4.9 |
| Age of deceased in years (SD) | 24.9 ± 9.1 |
| *Relationship to deceased* |  |
| Parent | 9 (50.0) |
| Sibling | 5 (27.8) |
| Partner | 4 (22.2) |
| *Clinical diagnosis of disease* |  |
| Inherited cardiomyopathy | 5 (27.8) |
| Inherited arrhythmia syndrome | 4 (22.2) |
| Unexplained | 9 (50.0) |
| *Variant identified* |  |
| LP/P variant | 7 (38.9) |
| VUS | 6 (33.3) |
| No variant | 5 (27.8) |
| *Reclassification* |  |
| Upgrade | 1 (5.6) |
| Downgrade | 2 (11.1) |
| No reclassification | 15 (83.3) |

Abbreviations: SCD, sudden cardiac death; LP, likely pathogenic; P, pathogenic; VUS, variant of uncertain significance; SD, standard deviation.

**FIGURE 1:**
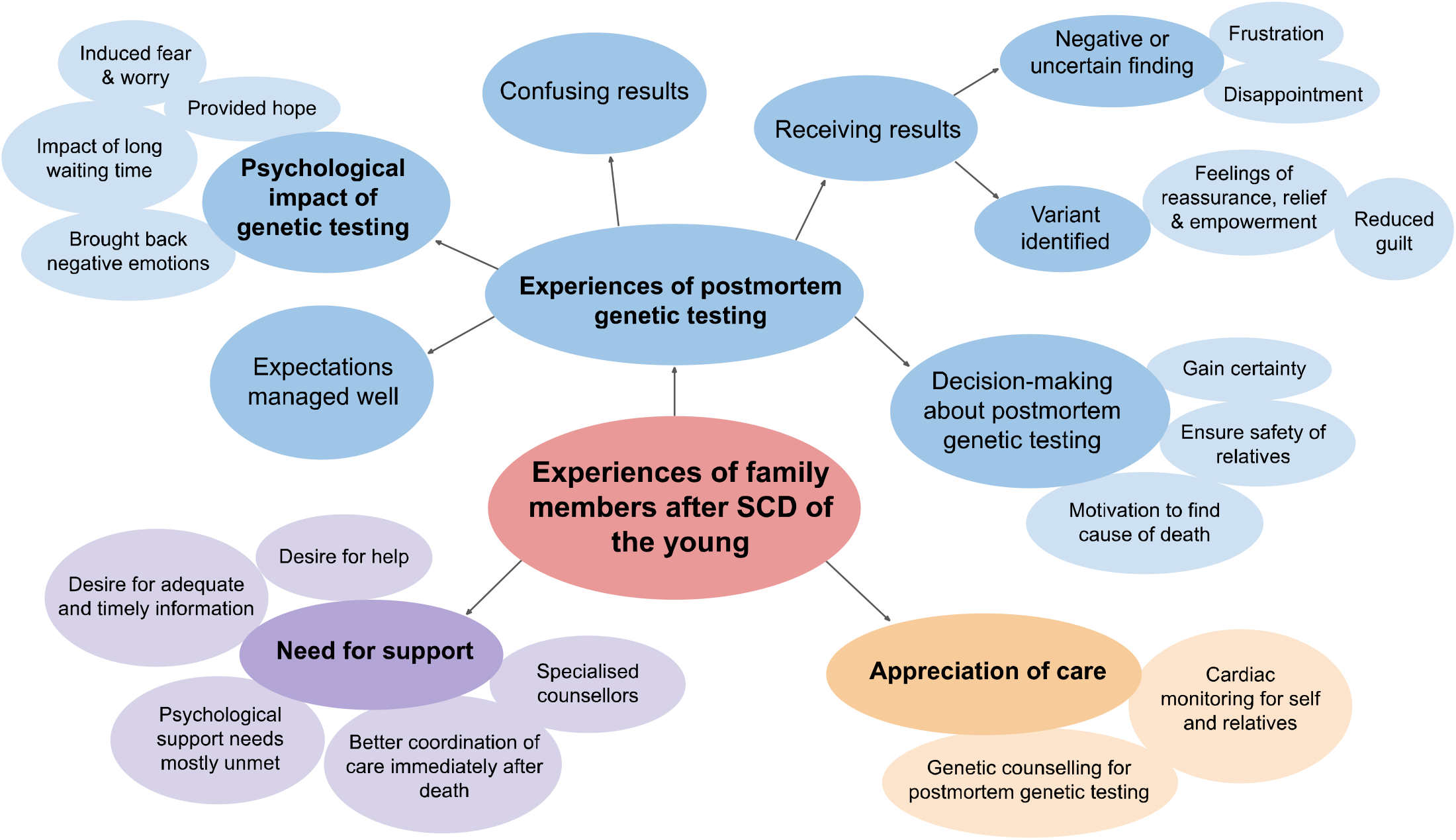
Summary of themes identified in the study. Abbreviation: SCD, sudden cardiac death.

### Experiences of family members

Based on thematic analyses, three main themes were identified: (1) Experiences with postmortem genetic testing, (2) appreciation of care, and (3) need for support.

#### Experiences with postmortem genetic testing

##### Decision-making about postmortem genetic testing

Participants’ main motivation for postmortem genetic testing was that it potentially provided an answer, a cause of death. Some mentioned that they wanted to do this to make sure they would not miss something and potentially gain certainty.

“*I just hope that one of these years maybe we’ll get an answer. I guess, again, as a mother, you want to know it wasn’t something you missed. That bothered me for quite some time and now I think well I cannot think of anything that we missed or could have done differently.”* (Participant 18, parent)

Most participants said that family members felt the same way about genetic testing. Further, participants decided to do it to ensure relatives are safe and to prevent sudden death from happening to others. Many participants said they had no reservations about postmortem genetic testing; it just felt like something logical to do.

“*I had no reservations at all about [genetic] testing. I couldn’t think of one reason why not.”* (Participant 10, sibling)

##### Receiving results of postmortem genetic testing

For participants where a causative variant was identified in the family, the result led to feelings of reassurance, relief and empowerment as it confirmed a course of action for clarifying risk to family members.

“*Knowing also that information has empowered us to ensure that I don’t pass it onto my children and that’s a massive part of the benefit we’ve gained from the genetic testing. It’s almost like a bit of gift going forward from my brother, that’s concern that he died from and that’s what I’ve got. Now it’s the positive thing about it.”* (Participant 17, sibling)

Some participants also mentioned that knowing the genetic cause diminished their feelings of guilt that they perhaps could have prevented the death from happening. For participants where postmortem genetic testing in the family was negative or identified an uncertain finding, postmortem genetic testing caused frustration and disappointment.

“*All these testing and genetic testing or whatever, they still can’t find it. Like that was – to my ears, I don’t know, all these tests and stuff you’d think something would come up or whatever, but nothing’s come up yet for him, I guess. [It’s] 2019, you know, and you still can’t find something.”* (Participant 11, sibling)

However, some also said that the cardiologist and genetic counsellor helped them manage their expectations well, by having told them prior to genetic testing that the chance of finding a cause was small. Understanding the result was complicated for some people, especially for participants with lower health literacy.

“*I say I think I would only have a very vague sense of what’s implied in that [genetics] and actually what – I have certain visuals in my head as to what might be involved, this sort of stupid vision, basic stupid visuals to with the DNA spirals or something like in the way in which things might operate in combinations, and then what might be applied in that, but I have no idea really.”* (Participant 12, parent)

##### Psychological impact of postmortem genetic testing

Participants experienced the impact of postmortem genetic testing differently. While for some it had a positive impact, others experienced negative impact. For many participants, postmortem genetic testing felt like an opportunity to get an answer.

“*It made me feel a little bit empowered that the more knowledge we have the better, even if it’s not the great news, we at least know. I’m very proactive in that regard so any opportunity to have a much more informed position of what’s going on genetically in our family”* (Participant 9, sibling)

Postmortem genetic testing therefore provided hope, which participants experienced as a positive impact. For some participants, it was hard to wait for results while desperately looking for answers about what happened, especially if they had to wait some time for the results of the postmortem investigation before that.

“*That wait of that couple of times [coroner, postmortem investigation report, getting appointment with specialized cardiologist] was pretty awful because we had questions that were unanswered and we went down and saw our GP and spoke to him about it He was able to diffuse what all that meant. But I guess it’s that timeframe from when it happened and then to actually get more information from a professional person that was tough.”* (Participant 13, parent)

For others, the waiting time was less of an issue, as there were many other things to arrange or because they felt more at peace with not knowing the cause of the death. Some participants said postmortem genetic testing brought back emotions surrounding the traumatic death, also because the results took a long time.

“*After a few months after that subsided, and because I hadn’t had any information, I just thought to myself “Oh well, we don’t know why it happened, we’ll never know why” and I think I did sort of not think any more about the whole genetic testing thing and it was literally out of the blue when I got the information back again. It was “Ah yeah that’s right. You guys are doing that testing”. I’d just forgotten about it.”* (Participant 16, partner)

Others mentioned they felt nervous about the genetic testing results and that it induced fear and worry, e.g., of getting a positive result, of implications of being affected, and of sharing results with parents and other family members. Many participants also said they thought it was hard to hear there are samples kept from the deceased, while they often were unaware of that during the postmortem investigation procedure.

“*I asked them* [coroner’s office] *if they had any samples and they came back and said yes, they gave us quite a list of all the different samples they had, like part of his lung and his heart, and this part of the heart. I guess it was like “oh so [name brother]’s still here”, because I didn’t know they had the samples and in my mind he’s dead and he’s gone but now they’re saying they’ve got a piece of him. So, to me that was a bit weird.”* (Participant 7, sibling)

#### Appreciation of care

All participants in this study were eventually seen in a specialised cardiac genetics clinic, where they received genetic counselling for postmortem genetic testing and cardiac monitoring for themselves and other family members. While all participants appreciated the care they received at the specialised clinic, many of them felt there was a lack of coordination of care in the preceding period (i.e., immediately after the death), leading to participants feeling left alone.

“*It was a long time of nothing to be honest. It was a long time of … There was no contact with anyone because no one knew how he [son] died, so if there is a way of keeping these families linked in just so that they know that there is an option going forward that if more information came to light or if another family member knew who to turn to. I think a lot of these families are just left in the dark because there’s no answer and there’s no certainty and unless you happen to have someone who has an understanding of these things then it would be easy to feel alone.”* (Participant 17, parent)

Some participants said that no one mentioned the possibility of postmortem genetic testing, that they were not referred to a cardiologist but had to navigate this on their own, and that there was a long waiting time to see a cardiologist for cardiac monitoring. Some felt there was also a lack of transparency about the sample that was taken during the postmortem examination, while participants or other family members were not asked to provide consent.

Some participants said they felt the information they received was good and appreciated that information provided was tailored to their needs. Other participants however felt there was a lack of or inconsistent information on several topics, including advice for relatives, genetic inheritance and testing, and updates on research. They felt they had to do their own research to find answers.

*“I suppose nobody still had definite answers and then sometimes you get to the answer and it might not be exactly that. Obviously I don’t have the scientific background to understand everything, so I was trying to Google and read up things that I didn’t understand. I just felt a bit muddled all the time. Try to take in any information and I think because I don’t come from, I really wanted to understand everything a bit, I don’t have a medical or scientific background, it was complex.”* (Participant 3, parent)

While support of specialized and knowledgeable healthcare professionals was positively evaluated, the psychological support needs for most participants were reported as unmet. Specifically, participants mentioned that counsellors (i.e., grief counsellors or community psychologists) did not have expertise in sudden death and that there seemed to be no advice regarding how to cope and deal with grief. They also experienced a long waiting time to see a counsellor and had to pay for psychosocial support. Some participants also experienced that their concerns about their surviving children were disregarded.

*“To me it’s remarkable for me that my – seeing a psychologist, it’s as if they’ve got no way of dealing with – or they’ve got no structure or thought pattern or way of working with somebody’s grief.”* (Participant 2, parent)

Some had eventually encountered positive experiences with a grief counsellor, including normalizing feelings, helping to relax and gain control, asking the right questions and being receptive.

Participants reported different experiences with the communication of healthcare professionals: Some were very empathic and sensitive, while other healthcare professionals were not. Insensitivity and lack of knowledge by healthcare professionals regarding the death and their feelings and emotions was by some participants experienced as very hurtful and burdening.

“*The* [employee] *at the Coroner’s office, when we were trying to get some answers, he said “Oh I understand exactly how you must be feeling it’s so frustrating. My wife died some years ago but eventually you’ll move on. I moved on and remarried and I have a new wife and everything is really good now”. I said “You can’t compare that. I’m in my 50s and I can’t have another son. It’s impossible. You can’t compare your wife dying to my son dying".”* (Participant 8, parent)

Knowledge or appreciation by healthcare professionals regarding grief was considered important. A few participants specifically mentioned they appreciated healthcare professionals who gave time to digest the information they received, especially while grieving. In addition, many participants also said they appreciated the possibility to contact the healthcare professional again if needed.

*“I think the things I really valued were they [HCPs] were really empathetic. They talked to us like we had some understanding of what was going on. They talk to me in a way that was respectful and they acknowledged that we – they would have a conversation with us anytime if we had questions about things.”* (Participant 4, parent)

#### Needs for support

##### Desire for adequate and timely information

Some participants specifically said they preferred to receive a timeline of what to expect, in order for family members to prepare themselves for emotional or confronting things that happen after the death.

*“If somebody had said at that point, and this probably as a general, if they said “look here’s the process of what’s going to happen here, we’re going to do this, body taken here, the Coroner”. If somebody said tissue samples, could be like complete organs. I don’t think it’d make it any better but it might be – when it comes back it would be less of a shock.”* (Participant 6, parent)

Also, the waiting time for genetic testing and results was burdensome for some, and therefore it was desired to have frequent updates of the genetic testing, even to just hear that testing is still in progress. Further, transparency about the sample taken during postmortem examination (for DNA extraction) of the deceased was also preferred by some participants. They mentioned that not knowing a sample was taken at postmortem examination and hearing that afterwards, was confronting and emotionally burdening. A desire for reliable information resources and information on whom to contact for help or support was also expressed by some participants.

##### Desire for help

The desire to know what happened to get closure about the death was significant for all participants. Therefore, better coordination of care for SCD families and early on contact with a specialized or knowledgeable healthcare professional were desired by some participants.

*“I think a bit of proactive medical and psychological – like if I didn’t go to the GP he would never have known about it. If I didn’t ask all the questions, and it is still my responsibility but I’m not in my clearest mind. Yeah, a bit of a case worker or something just to hold our hands a bit and give us some options and tell us what the government can do for us.”* (Participant 2, parent)

Many participants, especially parents, mentioned their main worry after the sudden death of their loved one was the potential risks for their remaining children and other family members. Therefore, some participants desired clear referral pathways to a specialized clinic for cardiac monitoring. To prevent SCD in other families, many participants also expressed the desire to participate in research or to help provide support to families who need that.

##### Desire for support

As previously described, support needs, especially psychosocial support, were expressed by many participants. Many participants wanted psychosocial support from professionals with expertise in SCD. A few participants also needed school counselling for their children or relational therapy for themselves and their partner. In addition, many participants wanted to share their experiences and feelings with other families who have also experienced young SCD.

*“We were sent to see psychologists and that, it was just an absolute waste of time. It was a horrendous experience at home and they tried a couple of times and it was just awful. Then we came up [to the city] and had the support group with this – it must have been a dozen families and so there were more mums and dads [who lost a child]… So it was like okay I’m not the only one.”* (Participant 5, parent)

These participants searched for peer support in their immediate environment, and also online. While some participants mentioned they appreciated the possibility to contact healthcare professionals when needed, a few participants also expressed the need for ongoing psychosocial support.

*“I think it should be a combination of things that people can go to or not. I think it could be a network initially with online sort of stuff with then someone organizing it and staying in touch with what people might sort of like. I think the patient information days is a good idea. I think sort of face-to-face little group about issues maybe from talks about particular issues, maybe it might be some of that could be clinical knowledge, some of that could be consumers talking about their experience of some issue. I think a mix of things would be really good.”* (Participant 4, parent)

## DISCUSSION

We conducted a qualitative study to explore experiences with cardiogenetic evaluation, including clinical screening, genetic testing and psychological support, following a young SCD. We show that participants positively appreciated the possibility of postmortem genetic testing. The option of postmortem genetic testing was perceived by bereaved family members in this study as a way to potentially get an answer, minimising the uncertainty and facilitating the grieving process. The process of cardiogenetic evaluation both positively and negatively affected the wellbeing of bereaved family members. On one hand it provided hope for getting an answer and ensuring the safety of family members; but it also raised uncertainty and led to a revival of emotions surrounding the traumatic death. Similarly, it has been previously reported that family members perceived this as a drawback of postmortem genetic testing (11).

In the current study, family members of SCD victims in whom no genetic cause was identified indicated that it was important that their expectations regarding outcomes of postmortem genetic testing were well-managed. It was easier to process the outcome as they knew there was a small chance a genetic variant would be identified. The role of genetic counselling in this process to adequately inform grieving family members and manage the expectations about potential outcomes is therefore critical (12,13). As stated in the APHRS/HRS expert consensus document, genetic counselling is integral to the process of cardiogenetic evaluation after SCD (8). Furthermore, in clinical practice, healthcare professionals involved should be cognizant of the impact that waiting times and uncertainty can have in this setting, and optimally support family members in this regard (9).

Almost all participants in our study experienced a lack of coordination of care, prior to the point of being referred to a specialised cardiac genetic clinic. Participants felt they had been left alone to find answers and ensure family members are cared for. Importantly, many participants felt they had to wait too long to get the specialized care needed. Similar findings have been reported and highlight the lack of coordination of care globally, with many navigating their way to specialised clinics using the internet and their own connections (6,11). Previous research suggests that access to multidisciplinary cardiac genetic care is limited and variably available worldwide, with knowledge of healthcare professionals also being a major barrier in individuals being referred to these services (14-16). This indicates a need to increase awareness among coroners and healthcare professionals of the need for specialized care after SCD and finding ways to increase access to specialized care after a young SCD (17,18).

Although the utility of cardiogenetic evaluation is widely recognized, there are concerns about how family members cope with this process of clinical, genetic and psychosocial evaluation (13). Many participants in this study expressed needs for adequate and timely information, sensitivity by professionals, and help to make sure that family members are safe and receive psychological care. The impact on psychological and family functioning is profound (5,19). Needs for psychological care and peer support of participants in this study were mostly unmet; many participants felt that the psychologists and grief counsellors they visited had no expertise regarding grief after SCD. Support networks were also limited. While needs were identified in many areas, a previous needs analysis study among suddenly bereaved family members (7) also showed that needs for psychological care were most often unmet. As suggested by Steffen et al (20), ongoing psychosocial support programs, including peer support, should therefore be integrated in clinical care for these families. Indeed, recent expert consensus guidelines support clinical psychological support for all family members following a young SCD (8).

This study had some limitations. For one interview, the quality of the recording affected our ability to transcribe the data. This interview was therefore excluded from the analysis. The time since the death differed between participants and lasted up to 20 years meaning participants had a wide range of experiences given evolution of genetic testing over this time. Recall of experiences surrounding the death and the period after that was therefore likely to be affected.

## CONCLUSION

The SCD of a young person has profound implications for the surviving family members. There are significant efforts worldwide to ensure families have access to clinical evaluation, genetic testing and psychological support. The impact of these investigations and support are overall positive, though there are important factors to consider including better coordination of care from the point of death to attendance at a specialised clinic, ensuring sensitivity of healthcare professionals and clear but timely provision of information.

## Data Availability

All data produced in the present study are available upon reasonable request to the authors

## ACKNOWLEDGEMENTS

We thank all participants for sharing their experiences with us. LY is a recipient of a co-funded National Heart Foundation of Australia/National Health Medical Research Council (NHMRC) PhD Scholarship (#102568/ #191351). CS is the recipient of an NHMRC Practitioner Fellowship (#1154992) and a New South Wales (NSW) Health Cardiovascular Health Clinician Scientist Grant. JI is the recipient of a NHMRC Career Development Fellowship (#1162929). This study was funded in part by a (NSW) Health Cardiovascular Research Capacity Program early-mid career research grant (JI).

## ETHICS APPROVAL

Sydney Local Health District (Royal Prince Alfred Hospital Zone) Ethics Committee gave ethical approval for this work (X19-0019 & 2019/ETH00094). Participants provided written, informed consent.

## REFERENCES

1. Bagnall RD, Weintraub RG, Ingles J et al. A Prospective Study of Sudden Cardiac Death among Children and Young Adults. N Engl J Med 2016;374:2441–52.

2. Lahrouchi N, Raju H, Lodder EM et al. Utility of Post-Mortem Genetic Testing in Cases of Sudden Arrhythmic Death Syndrome. J Am Coll Cardiol 2017;69:2134–2145.

3. Skinner JR, Crawford J, Smith W et al. Prospective, population-based long QT molecular autopsy study of postmortem negative sudden death in 1 to 40 year olds. Heart Rhythm 2011;8:412–9.

4. Yeates L, Hunt L, Saleh M, Semsarian C, Ingles J. Poor psychological wellbeing particularly in mothers following sudden cardiac death in the young. Eur J Cardiovasc Nurs 2013;12:484–91.

5. Ingles J, Spinks C, Yeates L, McGeechan K, Kasparian N, Semsarian C. Posttraumatic Stress and Prolonged Grief After the Sudden Cardiac Death of a Young Relative. JAMA Intern Med 2016;176:402–5.

6. Wisten A, Zingmark K. Supportive needs of parents confronted with sudden cardiac death--a qualitative study. Resuscitation 2007;74:68–74.

7. McDonald K, Sharpe L, Yeates L, Semsarian C, Ingles J. Needs analysis of parents following sudden cardiac death in the young. Open Heart 2020;7.

8. Stiles MK, Wilde AAM, Abrams DJ et al. 2020 APHRS/HRS expert consensus statement on the investigation of decedents with sudden unexplained death and patients with sudden cardiac arrest, and of their families. Heart Rhythm 2021;18:e1–e50.

9. Bates K, Sweeting J, Yeates L, McDonald K, Semsarian C, Ingles J. Psychological adaptation to molecular autopsy findings following sudden cardiac death in the young. Genet Med 2018.

10. Braun V, Clarke V. Using thematic analysis in psychology. Qualitative Research in Psychology 2006;3:35.

11. van der Werf C, Onderwater AT, van Langen IM, Smets EM. Experiences, considerations and emotions relating to cardiogenetic evaluation in relatives of young sudden cardiac death victims. Eur J Hum Genet 2014;22:192–6.

12. Grubic N, Puskas J, Phelan D, Fournier A, Martin LJ, Johri AM. Shock to the Heart: Psychosocial Implications and Applications of Sudden Cardiac Death in the Young. Curr Cardiol Rep 2020;22:168.

13. Ingles J, James C. Psychosocial care and cardiac genetic counseling following sudden cardiac death in the young. Progress in Pediatric Cardiology 2017;45:31–36.

14. van den Heuvel LM, Do J, Yeates L et al. Global approaches to cardiogenetic evaluation after sudden cardiac death in the young: A survey among health care professionals. Heart Rhythm 2021;18:1637–1644.

15. Liu G, MacLeod H, Webster G, McNally EM, O’Neill SM, Dellefave-Castillo L. Genetic Counselors’ Approach To Postmortem Genetic Testing After Sudden Death: An Exploratory Study. Acad Forensic Pathol 2018;8:738–751.

16. Michaud K, Mangin P, Elger BS. Genetic analysis of sudden cardiac death victims: a survey of current forensic autopsy practices. Int J Legal Med 2011;125:359–66.

17. Semsarian C, Ingles J, Wilde AA. Sudden cardiac death in the young: the molecular autopsy and a practical approach to surviving relatives. Eur Heart J 2015;36:1290–6.

18. Fleming PJ, Blair PS, Sidebotham PD, Hayler T. Investigating sudden unexpected deaths in infancy and childhood and caring for bereaved families: an integrated multiagency approach. Bmj 2004;328:331–4.

19. Mayer DD, Rosenfeld AG, Gilbert K. Lives forever changed: family bereavement experiences after sudden cardiac death. Appl Nurs Res 2013;26:168–73.

20. Steffen EM, Timotijevic L, Coyle A. A qualitative analysis of psychosocial needs and support impacts in families affected by young sudden cardiac death: The role of community and peer support. Eur J Cardiovasc Nurs 2020;19:681–690.

